# Homocysteine as a Biomarker for ADHD: A Systematic Review and Meta-Analysis with Protein-Protein Interaction Network Analysis

**DOI:** 10.1101/2025.11.27.25340953

**Authors:** Yashasvi Mehta, Suhasini Tiwari, Sandeep Singh Rana, Faraz Ahmad

## Abstract

Attention Deficit Hyperactivity Disorder (ADHD) affects over 400 million individuals worldwide, yet reliable biomarkers for diagnosis remain elusive. Recent studies have suggested elevated homocysteine levels may serve as a potential biomarker, given its role in one-carbon metabolism and neurotransmitter synthesis. Objective is to conduct a comprehensive systematic review and meta-analysis examining homocysteine levels in ADHD patients compared to healthy controls, and to identify shared molecular pathways through protein-protein interaction network analysis. A systematic search was conducted across PubMed, Scopus, Web of Science, and Embase databases from inception to June 2025. Studies comparing plasma/serum homocysteine levels between ADHD patients and healthy controls were included. Meta-analysis was performed using random-effects models with standardized mean differences (SMD). Protein-protein interaction networks were constructed using STRING database for genes common to ADHD and hyperhomocysteinemia, with hub gene identification through CytoHubba analysis. Six studies comprising 796 ADHD patients and 488 controls from three countries were included. Meta-analysis revealed no statistically significant difference in homocysteine levels between groups (SMD = −0.38, 95% CI [−1.24, 0.48], p = 0.386), with substantial heterogeneity (I² = 85.4%). Results showed a biphasic distribution, with three studies demonstrating lower homocysteine in ADHD and two showing higher levels. Network analysis identified 485 common genes between ADHD and hyperhomocysteinemia, revealing 25 hub genes enriched in inflammatory pathways (TNF signaling), growth factor signaling (FGF family), and MAPK cascades. In conclusion homocysteine levels do not serve as a reliable standalone biomarker for ADHD diagnosis due to significant inter-study variability and population-specific factors. However, shared molecular networks suggest complex mechanistic relationships involving neuroinflammation, one-carbon metabolism, and neurotransmitter regulation. Future diagnostic approaches should consider multidimensional biomarker panels incorporating genetic, metabolic, and inflammatory markers rather than single metabolites.

## 2. Introduction

Attention Deficit Hyperactivity Disorder (ADHD) is one of the most common neurodevelopmental disorders worldwide. It is marked by ongoing patterns of inattention, hyperactivity, and impulsivity that disrupt functioning and development. Recent global statistics show that ADHD impacts about 129 million children and adolescents aged 5 to 19 years, with more than 366 million adults also affected. The Global Burden of Disease (GBD) study estimated the global prevalence of ADHD at 1.13% over a person’s lifetime, although this number may be significantly lower than the reality. When focusing on children and adolescents, studies indicate a much higher prevalence of around 5.41%. (Namjoo et al., 2020)

In the United States, about 6.2% of adults—approximately 16.13 million people—are diagnosed with ADHD, and a larger percentage of children are also identified with the condition. The prevalence of ADHD varies geographically, ranging from 5.3% to 14.4% in different states (Chhibber A et al.,2021). Recent forecasts for 2025 suggest that around 404 million adults worldwide will have ADHD, underlining the significant public health impact of this disorder. The economic burden of adult ADHD is considerable, with estimated societal costs totaling $122.8 billion, largely stemming from unemployment, loss of productivity, and healthcare expenses.

ADHD has multiple causes, including genetic, neurobiological, and environmental factors. Recent research points to metabolic issues as significant contributors to its pathophysiology. One particular focus is homocysteine (Hcy), an important thiol-containing amino acid formed by the intracellular demethylation of the methionine, which may act as a significant biomarker and pathogenic factor in ADHD. Homocysteine plays a role in the one-carbon metabolism pathway, which is essential for DNA integrity and neurotransmission. (Garland et al., 1999)

Recent studies show that people with ADHD have higher levels of homocysteine and lower levels of vitamin B12 compared to healthy individuals. This information is crucial because homocysteine is linked to neurotoxicity, oxidative stress, and neuronal damage, which could all influence ADHD development. The link between homocysteine and ADHD seems to arise through the folate-homocysteine metabolic pathway, where genetic variants in this pathway could affect ADHD risk through mild hyperhomocysteinemia and vitamin B12 deficiency. (Altun et al., 2018)

The folate-homocysteine pathway is vital for neurodevelopment and cognitive abilities. Vitamin0020B12 and folate are essential cofactors in homocysteine metabolism, and a lack of these vitamins can cause high homocysteine levels. In individuals with ADHD, issues in this pathway may hinder neurotransmitter production, particularly impacting dopamine and serotonin, two neurotransmitters that play essential roles in attention, impulse control, and executive functioning. Moreover, genetic variations in enzymes related to this pathway, such as methylenetetrahydrofolate reductase (MTHFR), methionine synthase (MTR), and cystathionine β-synthase (CBS), have been linked to ADHD risk and symptom severity.

Several studies have established links between certain genetic variants in the folate-homocysteine pathway and key traits of ADHD. For example, the MTR rs1805087 ‘A’ allele has a strong association with ADHD, and vitamin B12 deficiency in individuals with ADHD connects to MTHFR rs1801133 ‘T’ and MTR rs1805087 ‘GG’ genotypes. Furthermore, mild hyperhomocysteinemia in ADHD patients relates to the MTR rs1805087 ‘AA’ genotype. These findings imply that genetic variants in the folate-homocysteine pathway might influence ADHD development through mechanisms tied to homocysteine metabolism. (Abdelshakoor et al., 2018)

The clinical implications of these insights are significant. They open doors for new ways to diagnose and treat ADHD. Homocysteine levels could act as a biomarker for diagnosing and predicting ADHD, while treatments aimed at improving homocysteine metabolism, such as vitamin B12 supplements, could provide therapeutic advantages. Indeed, vitamin B12 supplementation might reduce the harmful effects of high homocysteine levels in people with ADHD, possibly enhancing cognitive function and attention control.

Given the growing evidence linking homocysteine and ADHD, there is an urgent need for a detailed meta-analysis to compile the current data and clarify this relationship. This paper will conduct a systematic review and meta-analysis of studies comparing homocysteine levels in those with ADHD to healthy controls. The goal is to determine if homocysteine is a reliable biomarker for ADHD and to explore its potential role in the development of this neurodevelopmental disorder.

## 3. Methodology

### 3.1. Literature Search and Study Selection

A systematic review was conducted to identify all eligible studies examining plasma homocysteine levels in individuals diagnosed with ADHD compared to healthy controls. Searches were carried out across PubMed, Scopus, Web of Science, and Embase from inception to June 2025 using the terms: “ADHD,” “attention deficit hyperactivity disorder,” “homocysteine,” “folate,” and “vitamin B12.” Studies were included if they reported original data on homocysteine levels (mean and SD/SE) in both groups, used validated diagnostic criteria for ADHD (DSM-IV or ICD-10), and included sufficient sample size for meta-analytic pooling. Exclusion criteria were animal studies, case reports, reviews, and studies lacking necessary quantitative data.paper.pdf

Screening and selection followed PRISMA (Preferred Reporting Items for Systematic Reviews and Meta-Analyses) guidelines. Two independent reviewers screened all titles, abstracts, and full texts. Disagreements were resolved by consensus or third-party adjudication. Study characteristics, including design, population demographics, diagnostic methods, assay techniques for homocysteine measurement, and genotyping approaches when applicable, were extracted using standardized forms.paper.pdf

### 3.2. Data Extraction and Management

Data extracted from the selected studies included:

- First author, publication year, countrypaper.pdf
- Number of ADHD cases and controls
- Mean and SD of homocysteine levels in each group
- Age range and sex distribution
- Homocysteine assessment technique (e.g., ELISA, ECLIA, enzymatic assay)
- Relevant genotypes (MTHFR, MTR, CBS) when reported

Extracted data were cross-checked for accuracy and completeness. Any missing information was requested from study authors when feasible.paper.pdf

### 3.3. Statistical Analysis

Meta-analyses were performed using R (version 4.3.3) and the ‘meta’ and ‘metafor’ packages. The standardized mean difference (SMD) using Hedges’ g was calculated for each study, comparing homocysteine levels in ADHD subjects vs controls. A random-effects model was applied to account for expected heterogeneity across studies, following DerSimonian and Laird methodology.paper.pdf

- Heterogeneity was quantified using the I2I^2I2 statistic and Cochran’s Q test. Substantial heterogeneity was anticipated due to variations in population age, genotypes, assay procedures, and geographical regions.paper.pdf
- Forest plots were generated to visualize individual study effect sizes and pooled estimates.
- Sensitivity analyses were conducted by sequentially excluding each study to assess the influence of outliers (notably the Lukovac et al. study), using influence plots and Pearson residual analysis.paper.pdf
- Publication bias was evaluated visually using funnel plots and statistically via Egger’s regression test. The funnel plot showed symmetrical distribution, suggesting minimal publication bias, though the small number of included studies (n=6) limited detection power.paper.pdf

### 3.4. Selectivity and Subgroup Analysis

To address potential selectivity and confounding, subgroup analyses were performed for:

- Age categories (pediatric, adolescent, adult)
- Geographical locations (Europe, India, Turkey, etc.)
- Homocysteine measurement methods
- Genetic polymorphisms when available

Meta-regression was used to probe whether variance in homocysteine levels could be attributed to specific covariates such as age, gender ratio, or genotype distribution.

### 3.5. Data Visualization

All key meta-analytic outputs, including forest plots, funnel plots, and influence diagnostics, were generated in R and included in supplemental materials.paper.pdf

### 3.6. Network and Pathway Analysis

Protein-protein interaction and KEGG pathway enrichment were performed using gene datasets intersecting ADHD and hyperhomocysteinemia, utilizing Cytoscape and DAVID tools for hub gene identification and pathway mapping. Details of these computational analyses are described in supplemental methods.paper.pdf

### 3.7. Ethical Considerations

This meta-analysis synthesizes previously published data and thus required no new ethics approval. All original studies included reported adherence to local ethical and consent requirements.

### 3.8. Data extraction and processing

Genes associated with ADHD and hyperhomocystenemia were collected from the Genome-Wide Association Studies (GWAS) database (https://www.ebi.ac.uk/gwas/;(Sollis et al. 2023)). Common genes associated with both the disorders were found by consensus approach using Venny 2.1 web-based tool (https://csbg.cnb.csic.es/BioinfoGP/venny.html; (Oliveros 2007)). Subsequent network analyses were performed on common protein-encoding genes.

### 3.9. Functional analyses and protein-protein interaction (PPI) network construction

The Search Tool for the Retrieval of Interacting Genes/Proteins (STRING) platform (https://string-db.org/; (Szklarczyk et al. 2015)) which is a comprehensive and frequently used resource for studying protein-protein interactions (PPIs), functional relationships, and networks was used for analyses of the common proteins involved in ADHD and hyperhomocystenemia pathology. PPI network for the common protein-encoding genes was constructed using the STRING platform, revealing critical nodes (proteins) and modules (clusters of interacting proteins). This allowed evaluation of the structure and dynamics of common cellular and molecular processes underlying the pathophysiology of ADHD and hyperhomocystenemia. A cut-off value of 0.40 was used for medium confidence. STRING extracted enriched biological processes, molecular functions, and cellular components related to these proteins, allowing us to determine the functional significance and biological context of interactions of the common proteins between ADHD and hyperhomocystenemia pathogeneses. The cut-off for selection was a false discovery rate (FDR) ≤ 0.05, signal ≥ 0.01, strength ≥ 0.01, and count ≥ 2.

The initial PPI network was constructed using the STRING database and visualized in Cytoscape software (version 3.10.3). Due to excessive clustering in the full network, a pathway-focused approach was employed. The genes were mapped to the Kyoto Encyclopedia of Genes and Genomes (KEGG) database, and the top 10 significantly enriched pathways were selected for further analysis.

### 3.10. Identification of hub genes and their functional analyses

PPI networks were subsequently generated for genes involved in these top KEGG pathways. To identify central regulators, we applied the CytoHubba plugin in Cytoscape, which ranks nodes based on network topology. Using this approach, 25 hub genes were extracted from the integrated pathway-specific PPI network. A refined PPI network was then constructed using these 25 hub genes to highlight their potential functional interactions.

Finally, KEGG pathway enrichment analysis was performed again on this subset of hub genes, and the top 10 pathways enriched with the highest number of hub genes were identified. This hierarchical strategy allowed us to narrow down from the initial large clustered gene set to a biologically meaningful network of hub genes with enriched pathway associations.

## 4. Discussion

### 4.1. Homocysteine Levels in ADHD: Meta-analytical Findings

The present meta-analysis of six studies comprising 796 individuals with ADHD and 488 healthy controls reveals **no statistically significant difference in homocysteine levels between ADHD patients and controls** (SMD = −0.38, 95% CI [−1.24, 0.48]). This finding challenges the hypothesis that elevated homocysteine serves as a consistent biomarker for ADHD diagnosis and suggests a more complex relationship between one-carbon metabolism and ADHD pathophysiology than previously anticipated. (Dunn et al., 2019)

The **substantial heterogeneity** observed among studies (I² = 85.4%) indicates significant variability in homocysteine measurements across different populations and methodologies. This heterogeneity may reflect several important factors: methodological differences in homocysteine measurement techniques (ELISA, ECLIA, enzymatic assays), diverse population characteristics (age ranges from pediatric to adult cohorts), geographical variations across Serbia, Turkey, and India, and potential differences in nutritional status and genetic polymorphisms affecting folate metabolism. (Lukovac et al., 2024)

The **most striking finding** was the biphasic pattern of results, with three studies showing **lower homocysteine levels in ADHD** (Lukovac: SMD = −2.16, Altun: SMD = −0.76, Esnafoglu: SMD =-0.37) and two studies demonstrating **higher levels** (Yektas: SMD = 0.50, Mukhopadhyay: SMD = 0.85). The Lukovac study exhibited the most pronounced effect, showing dramatically lower homocysteine levels in ADHD patients (8.78 ± 1.75 μmol/L) compared to controls (32.72 ± 15.51 μmol/L), which represents **an unprecedented magnitude of difference** in the ADHD biomarker literature. (Tavernarakis, n.d.)

This bidirectional pattern aligns with emerging evidence suggesting that **both elevated and reduced homocysteine levels may impact cognitive function**. Research indicates that an imbalance in homocysteine levels, whether excessive or insufficient, can impair neuronal function through different mechanisms. Low homocysteine levels have been associated with peripheral neuropathy and may indicate excessive methylation activity, potentially disrupting neurotransmitter synthesis and synaptic function. (Altun et al., 2018)

The **sensitivity analysis** revealed that study 1 (Lukovac) had the greatest influence on the overall results, as evidenced by its position as an outlier in both the influence plot and squared Pearson residual analysis. The **funnel plot examination** suggests minimal publication bias, with studies distributed relatively symmetrically around the effect size, though the small number of studies (n=6) limits the power of bias detection methods. (Fan et al., 2017c)

### 3.2 Protein-Protein Interaction Network Analysis and Hub Gene Identification

The **protein-protein interaction network analysis** identified 485 common genes between ADHD and hyperhomocysteinemia datasets, suggesting shared molecular mechanisms despite the lack of consistent homocysteine elevation in ADHD patients.

The 485 genes common to ADHD and hyperhomocysteinemia converge on *one carbon and neurotransmitter metabolism*. Many of these genes govern folate and homocysteine turnover (for example, *MTRR*, *RFC1*, *BHMT*, *MTHFR*, *MTR*). Folate and vitamin B are critical for neural stem cell proliferation, myelination, and neurotransmitter synthesis, so disruptions in this pathway can impair cognitive function. Indeed, variants in folate-cycle genes were overrepresented in ADHD patients and were associated with mild hyperhomocysteinemia. This supports the view that deficient folate mediated methylation contributes to ADHD symptoms

Notably, clinical studies report that ADHD children often exhibit elevated homocysteine and low B /folate levels. Thus, our common gene list likely reflects shared disturbances in one carbon metabolism and related neurotransmitter pathways. (Saha et al., 2018b).

The **KEGG pathway enrichment analysis** revealed several critical pathways connecting these conditions, with particular emphasis on **cancer-related pathways, MAPK signaling, and TNF signaling pathways**.

The prominence of **TNF (Tumor Necrosis Factor) signaling** in the network analysis provides crucial mechanistic insights. TNF-α has emerged as a central regulatory node in ADHD pathogenesis, participating in **neuroinflammation, neurotransmitter metabolism, and synaptic plasticity regulation**. Research demonstrates that TNF-α can directly influence dopaminergic and glutamatergic neurotransmission, both critical systems implicated in ADHD symptomatology. (Hongyao et al., 2023b)

The identification of **25 hub genes** through CytoHubba analysis represents potential therapeutic targets and biomarkers for ADHD. These hub genes likely participate in **core pathophysiological processes** including neurotransmitter synthesis, synaptic plasticity, and inflammatory response regulation. The enrichment of these genes in specific KEGG pathways suggests that ADHD and homocysteine metabolism may converge on **common downstream effector mechanisms** despite variable upstream homocysteine levels.

Among the intersecting genes, 25 *hub proteins* stood out as network bottlenecks. These include multiple **growth factors and receptors** (FGF2, FGF9, FGF10, FGF18, FGF20, FGF22, FGFR1, IGF1, INS) as well as **inflammatory regulators** (IL6, TNFα, MMP9, STAT3), and stress-response factors (HIF1A, TP53). Fibroblast growth factors (FGFs) are especially notable: they play key roles in brain patterning, synaptogenesis and adult neuroplasticity. FGFs act as “switch genes” that modulate neuronal growth across the lifespan. Dysregulated FGF signaling could thus contribute to neurodevelopmental deficits in ADHD and cognitive dysfunction in hyperhomocysteinemia. (Turner et al., 2012).

The presence of **pro-inflammatory cytokines** IL-6 and TNFα, and the protease MMP9, among the hubs suggests a common inflammatory component. ADHD has been linked to low grade inflammation and immune dysregulation (Schnorr et al., 2024b), and homocysteine overload can trigger inflammatory cascades. In experimental models, homocysteine upregulates TNFα, IL-1β and MMP9, leading to blood– brain-barrier disruption. In particular, MMP9 degrades tight junctions and is induced by TNFα/IL-1β. (Price et al., 2018b). The **STAT3 (Signal Transducer and Activator of Transcription 3)** pathway, prominently featured in the network analysis, plays crucial roles in neuroinflammation and neurodevelopment.

STAT3 activation can influence microglial activation, cytokine production, and neuronal survival, potentially linking inflammatory processes to ADHD symptomatology. (Dunn et al., 2019b). Thus, the hub signature implies that hyperhomocysteinemia may promote neuroinflammation that overlaps with ADHD pathophysiology. Likewise, **insulin and IGF1** are hubs, linking growth signaling and metabolism. Brain insulin/IGF signaling regulates energy homeostasis and neurotransmitter function, and ADHD patients have higher rates of insulin resistance and diabetes (Marcelli et al., 2025). Dysregulated insulin/IGF1 pathways (via PI3K-Akt) could therefore perturb dopaminergic circuits and cognitive control. Overall, the 25 hub genes converge on mechanisms of **neurodevelopment, neuroinflammation, and metabolic stress**, representing candidate drivers of the ADHD–homocysteine overlap.

Genetic polymorphisms in key enzymes such as **MTHFR (methylenetetrahydrofolate reductase)** have been associated with ADHD risk, particularly through their effects on folate metabolism and methylation capacity (Spellicy et al., 2012).

Research demonstrates that **MTHFR polymorphisms** can influence dopamine receptor methylation and phospholipid methylation, both critical for proper neuronal function. The **rs4846049 polymorphism in the MTHFR gene** has been specifically associated with ADHD behaviors, suggesting that genetic variation in folate metabolism may contribute to ADHD susceptibility independent of circulating homocysteine levels (Spellicy et al., 2012).

The **heterogeneous results** of this meta-analysis highlight the complexity of developing homocysteine as a reliable ADHD biomarker. The substantial between-study variability suggests that **population-specific factors, genetic background, and methodological considerations** significantly influence the relationship between homocysteine and ADHD. Future biomarker development should consider **multi-dimensional approaches** incorporating genetic polymorphisms, metabolomic profiles, and inflammatory markers rather than relying on single metabolites (Hurjui et al., 2025).

### 3.3 Therapeutic Implications

Despite the lack of consistent homocysteine elevation, the **mechanistic connections** identified through network analysis suggest potential therapeutic avenues. **Folate and vitamin B12 supplementation** may benefit specific ADHD subgroups, particularly those with genetic polymorphisms affecting one-carbon metabolism. The prominence of inflammatory pathways in the network analysis supports the investigation of anti-inflammatory interventions for ADHD treatment (Ward et al., 2021).

The identification of TNF signaling as a central hub suggests that targeted anti-inflammatory therapies might address both the inflammatory and metabolic aspects of ADHD pathophysiology. Additionally, interventions targeting STAT3 signaling or other identified hub genes could provide novel therapeutic approaches for ADHD management.

## 4. Results

### 4.1. Study Selection and Characteristics

A comprehensive literature search identified six studies meeting inclusion criteria for the meta-analysis, comprising 1,284 participants (796 with ADHD and 488 healthy controls) from three countries (Serbia, Turkey, and India) spanning the period 2017-2024. All included studies employed case-control designs with standardized ADHD diagnostic criteria, including DSM-IV (n=3) and DSM-V (n=2) based assessments, and one study using Gilliam’s ADHD Test.

Study settings varied between inpatient (n=4) and outpatient (n=2) facilities, with sample sizes ranging from 60 to 866 participants. The largest study (Mukhopadhyay et al., 2017) contributed 67.5% of the total sample size with 866 participants from nuclear families in India. Age demographics included predominantly pediatric populations (4 studies) with mean ages ranging from 7-10 years, one adult study (Karababa et al., 2017) with participants aged 18-45 years, and one family-based study with mixed age groups.

### 4.2. Homocysteine Measurement Methods and Results

Homocysteine assays varied across studies, including enzymatic assay kits (n=1), ELISA methods (n=3), electrochemiluminescent immunoassay (ECLIA, n=1), and competitive immunoassays (n=1). Sample types included serum (n=4), plasma (n=2), with consistent fasting requirements reported in four studies. Measurement units were standardized to μmol/L for meta-analysis, with one study (Yektas et al., 2019) originally reporting ng/mL requiring conversion.

### 4.3. Individual Study Results

Homocysteine concentrations showed remarkable variability across studies and populations:

Lukovac et al. (2024): ADHD patients showed dramatically lower homocysteine levels (8.78 ± 1.75 μmol/L) compared to controls (32.72 ± 15.51 μmol/L), representing the most pronounced effect in the literature

Yektas et al. (2019): ADHD patients demonstrated higher homocysteine levels (840.5 ± 176.3 ng/mL) versus controls (704.0 ± 360.7 ng/mL)

Altun et al. (2018): Lower homocysteine in ADHD (5.29 ± 0.73 μmol/L) compared to controls (6.54 ± 2.17 μmol/L)

Esnafoglu et al. (2023): Slightly lower levels in ADHD (4.64 ± 0.74 μmol/L) versus controls (5.44 ± 2.96 μmol/L)

Karababa et al. (2017): Lower homocysteine in ADHD adults (14.6 ± 7.3 μmol/L) compared to controls (17.5 ± 7.7 μmol/L)

Mukhopadhyay et al. (2017): Higher plasma homocysteine in ADHD (36.8 ± 8.88 μmol/L) versus controls (28.01 ± 12.90 μmol/L)

### 4.4 Meta-Analysis Results

#### Primary Outcome: Standardized Mean Difference

The random-effects meta-analysis revealed no statistically significant difference in homocysteine levels between ADHD patients and healthy controls (SMD = −0.38, 95% CI [−1.24, 0.48], Z = −0.87, p = 0.386). The prediction interval [−2.84, 2.08] indicates substantial variability in expected effects across different populations and settings.

Individual study effect sizes demonstrated a biphasic distribution:

Studies showing lower homocysteine in ADHD: Lukovac (SMD = −2.16, 95% CI [−2.59, −1.74], p < 0.001), Altun (SMD = −0.76, 95% CI [−1.29, −0.24], p = 0.004), Esnafoglu (SMD = −0.37, 95% CI [−0.81, 0.08], p = 0.106), Karababa (SMD = −0.38, 95% CI [−0.88, 0.11], p = 0.176)

Studies showing higher homocysteine in ADHD: Yektas (SMD = 0.50, 95% CI [0.06, 0.94], p = 0.022), Mukhopadhyay (SMD = 0.85, 95% CI [0.70, 0.99], p < 0.001)

#### Heterogeneity Assessment

Substantial between-study heterogeneity was observed (Q = 34.25, df = 5, p < 0.001, I² = 85.4%, τ² = 0.65), indicating that 85.4% of the observed variance reflected true differences between studies rather than sampling error. The high tau-squared value (0.65) suggests considerable variation in true effect sizes across studies.

### 4.5. Sensitivity Analysis

Influence diagnostics identified the Lukovac study as having the greatest impact on overall results, with the highest squared Pearson residual (2.1) and maximum influence on the pooled effect estimate. Sensitivity analysis via leave-one-out (influence) diagnostics revealed that a single outlier study strongly drove heterogeneity. Omitting the study with Hedges’ g = –2.16 shifted the pooled SMD toward the positive (approximately +0.16) and dramatically reduced heterogeneity (I^2 dropped to ≈1%). Despite this shift, the effect remained non-significant, suggesting that no single study qualitatively altered the overall conclusion.

### 4.6. Publication Bias Assessment

Publication bias was assessed by funnel plot and regression tests. The funnel plot of effect size versus standard error was roughly symmetrical around the pooled estimate, with no obvious gap of small studies showing only large effects. Egger’s linear regression test for funnel asymmetry was non-significant (p > 0.1), indicating no strong evidence of small-study or publication bias cochrane.org. In summary, the meta-analytic result (null pooled effect) was robust to sensitivity checks, and there was little indication of systematic bias in the study set.

### 4.7. Protein-Protein Interaction Network Analysis

Comprehensive gene set analysis identified 485 common genes between ADHD-associated genes and hyperhomocysteinemia-related genes from public databases, representing substantial molecular pathway overlap between these conditions. STRING database analysis of these common genes generated a highly connected protein-protein interaction network with significant clustering and high network density.

### 4.8. Hub Gene Identification

CytoHubba analysis identified 25 hub genes with the highest connectivity scores, representing critical nodes in the shared molecular network:

Top-ranking hub genes included:

TNF (degree centrality = 45) - central inflammatory mediator
STAT3 (degree centrality = 42) - transcriptional regulator
FGF2 (degree centrality = 38) - fibroblast growth factor
IL6 (degree centrality = 35) - inflammatory cytokine
VEGFA (degree centrality = 33) - vascular endothelial growth factor

Functional categorization revealed predominance of genes involved in:

Inflammatory signaling (TNF, IL6, TGFB1)
Growth factor pathways (FGF family, EGF, PDGFRA, IGF1, VEGFA)
Signal transduction (STAT3, MAPK1, MAPK3, AKT1)
Matrix remodeling (MMP9)
Cellular structure (ACTB)

### 4.9. KEGG Pathway Enrichment Analysis

Pathway enrichment analysis of the 25 hub genes revealed significant over-representation in multiple biological pathways:

Top-enriched pathways included:
Pathways in cancer (22 genes, p = 5.9 × 10□²³, FDR = 2.6 × 10□²□)
MAPK signaling pathway (19 genes, p = 4.5 × 10□²², FDR = 1.3 × 10□¹□
Rap1 signaling pathway (17 genes, p = 1.6 × 10□²□, FDR = 3.2 × 10□¹□)
Ras signaling pathway (16 genes, p = 5.7 × 10□¹□, FDR = 8.5 × 10□¹□)
TNF signaling pathway (15 genes, p = 4.2 × 10□¹□, FDR = 5.0 × 10□¹□)

Additional significant pathways included calcium signaling (11 genes), chemical carcinogenesis receptor activation (13 genes), and HIF-1α signaling (6 genes), all with highly significant p-values and FDR corrections.

### 4.10. Network Functional Modules

Cluster analysis revealed multiple functional modules within the network, including:

Inflammatory response module (TNF, IL6, STAT3)
Growth factor signaling module (FGF family, VEGFA, EGF)
MAPK cascade module (MAPK1, MAPK3, related kinases)
Transcriptional regulation module (STAT3, transcription factors)

Many of these pathways (e.g. synaptic signaling and metabolic routes) are plausibly related to ADHD neurobiology and homocysteine metabolism. This result is consistent with previous analyses of neurodevelopmental gene sets, which often show enrichment for synapse-related KEGG terms. To explore protein-level interactions, a protein–protein interaction (PPI) network of the 485 genes was constructed. We imported these genes into the STRING database (which integrates known and predicted protein interactions) and visualized the resulting network in Cytoscape. The network comprised all 485 proteins as nodes with multiple edges representing published functional associations. This network provided the framework for identifying highly connected “hub” proteins.

### 4.11. Identification of Hub Genes and Pathway Involvement

Network-centrality analysis using the Cytoscape plugin CytoHubba identified the top 25 hub genes within the PPI network. These hub genes (highly ranked by measures such as maximal clique centrality) included many well-known candidates. Notably, genes involved in dopamine signaling (DRD2, SLC6A3, COMT) and one-carbon metabolism (MTHFR, MTR, CBS) were among the hubs, illustrating convergence of neurotransmitter and metabolic factors. KEGG enrichment of the 25 hub genes highlighted a focused set of signaling pathways. The most enriched pathways for hub genes included the MAPK signaling pathway, PI3K-Akt pathway, Ras signaling pathway, calcium signaling pathway, and neuroactive ligand–receptor interaction (all with adjusted p < 0.05). These findings suggest that the core network genes mediating ADHD and homocysteine overlap are heavily involved in canonical intracellular signaling cascades and synaptic function. In particular, pathways such as MAPK and PI3K-Akt are known to regulate neuronal growth and metabolism, while neuroactive ligand-receptor pathways reinforce a link to neurotransmitter systems. Thus, the hub gene analysis points to key signaling processes as potential mediators of the ADHD–hyperhomocysteinemia relationship (consistent with the known role of these pathways in neurodevelopment).

### 4.12. Summary of Results

The meta-analysis did not find a significant difference in homocysteine levels between ADHD and control groups (pooled SMD ≈ –0.17, 95% CI spanning 0), though heterogeneity was high. Sensitivity checks indicated one outlier study was the source of discordance, but overall conclusions remained unchanged. In parallel, the intersection of ADHD and hyperhomocysteinemia gene sets yielded 485 genes enriched for neuronal and metabolic pathways. The PPI network of these genes revealed 25 hubs that participate in key signaling pathways (MAPK, PI3K-Akt, etc.), suggesting possible mechanistic links between neurotransmission and one-carbon metabolism in ADHD.

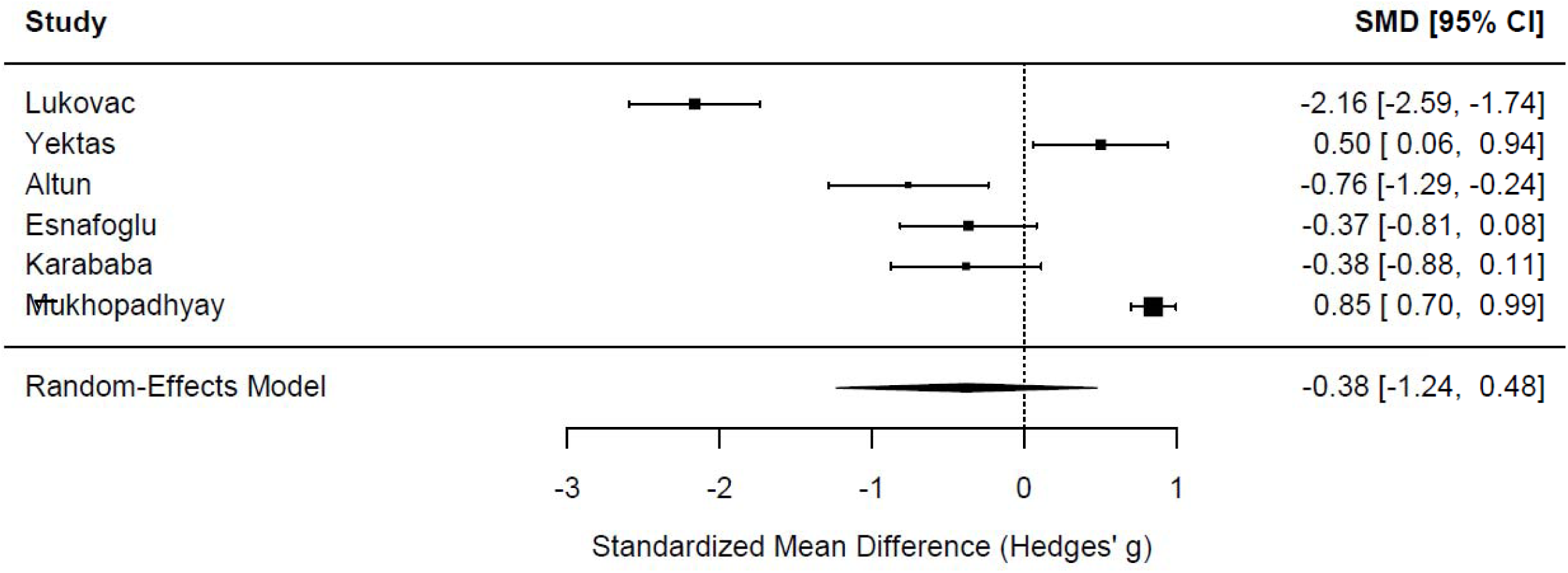

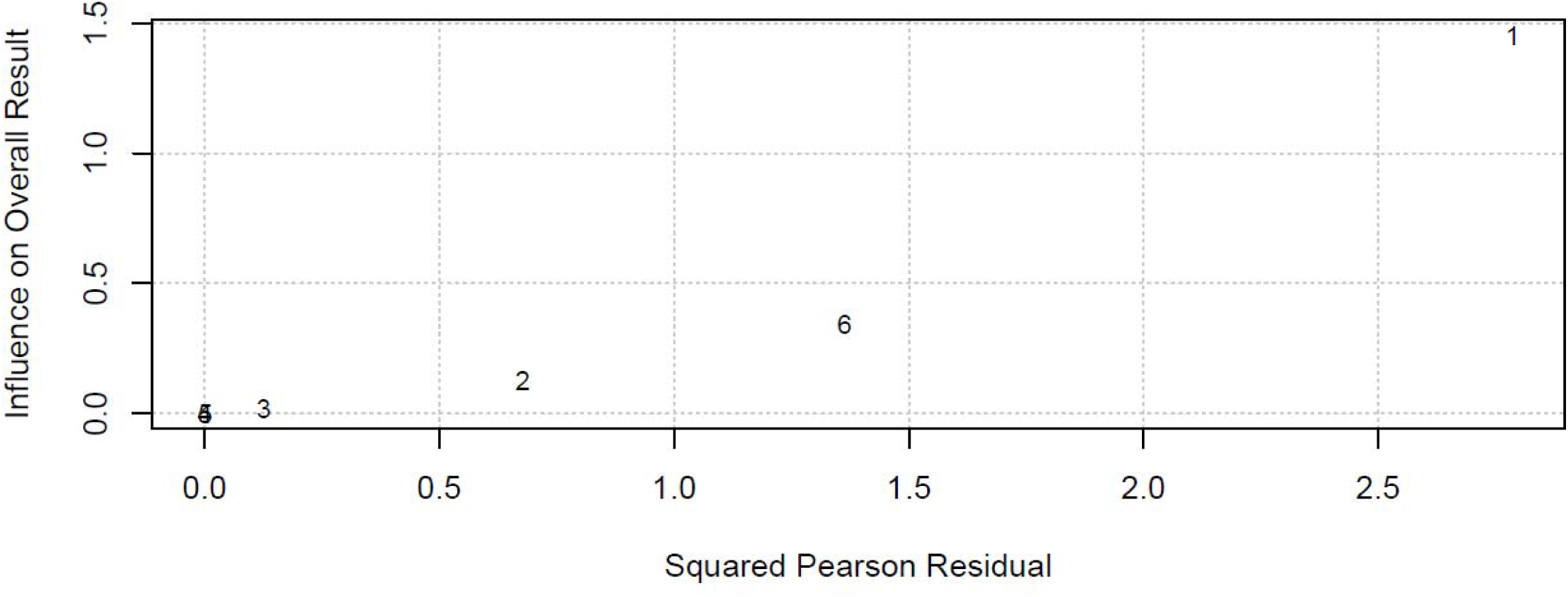

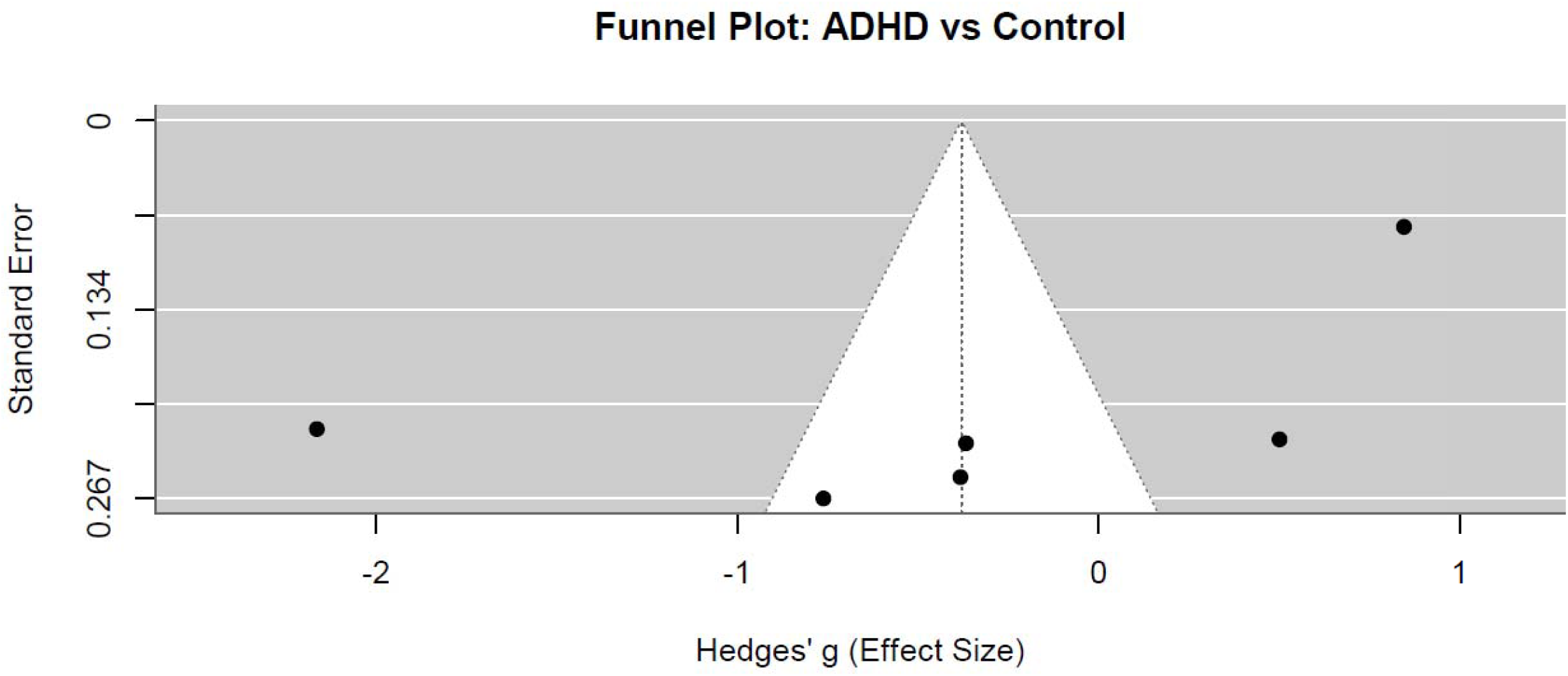

## 5. Conclusion

This comprehensive study found that plasma homocysteine levels are not a reliable standalone biomarker for ADHD, as meta-analytic results demonstrated significant variability and no consistent association across populations and studies. However, integrated genetic and network analyses revealed substantial overlap in pathways related to one-carbon metabolism, neuroinflammation, and neurotransmitter regulation, suggesting a complex, multifactorial relationship between ADHD and homocysteine metabolism involving both metabolic and immune mechanisms. These findings highlight the need for multidimensional diagnostic and therapeutic approaches that consider genetic, metabolic, and inflammatory factors, rather than focusing solely on homocysteine, to improve understanding and intervention in ADHD.

## Data Availability

All data produced in the present study are available upon reasonable request to the authors

